# Mobile money vendors as a community channel for adolescent condom provision and sexual health information in Uganda: a type-II hybrid effectiveness-implementation study

**DOI:** 10.64898/2026.11.15.26367221

**Authors:** Makiko Komasawa, Miho Sato, Sumihisa Honda, Paul Kitakule, Peter Waiswa

## Abstract

**Purpose:** Unmarried adolescents in Uganda have limited access to sexual and reproductive health (SRH) services, and conventional programs have struggled to reach them sustainably. Mobile money vendors (MMVs) are ubiquitous in young people’s daily lives. We assessed the feasibility of an MMV-based intervention to provide condoms and SRH information to adolescents.

**Methods:** In this type-II hybrid effectiveness-implementation study in Eastern Uganda, 30 MMVs provided oral and printed information, free condoms, and referrals for four months in 2024. Feasibility was assessed using the Bowen framework, drawing on service logs, baseline and endline surveys of adolescents aged 15–19 years (n = 1,204), three focus group discussions with MMVs, eight key informant interviews, a post-intervention MMV survey, and supervision reports.

**Results:** MMVs recorded 104,849 adolescent customer visits; oral information was provided at 54,365 visits and condoms at 21,056 visits. Condom use in the intervention arm increased from 67.3% to 77.7% (p = 0.04), and the proportion obtaining condoms from MMVs rose from 0% to 64.6%. Adolescents valued the service’s convenience and confidentiality. Initial resistance from some community adults largely diminished, although referrals remained few (210 cases). All MMV shops completed the intervention without disrupting business, and customer numbers and sales rose 1.64- and 1.50-fold. Suggested adaptations included vendor identification and services for girls, integration with public health systems for scalability.

**Conclusions:** The MMV-based approach was feasible and reached hard-to-access unmarried adolescents with condoms and SRH information. For scale-up, improvements for adaptation and large-scale evaluation are necessary.

**Trial registration:** UMIN000053332

## Introduction

Adolescent pregnancy is a global health issue that adversely affects birth outcomes and can lead to intergenerational cycles of poverty and ill-health [1 2]. Uganda faces these challenges acutely [3], with a notably young population—52.6% under 18 years [4]. The average ages of first marriage (18.7 years) and first sexual intercourse (16.9 years) among women 25–49 indicate that many girls are sexually active well before marriage [5]. Modern contraceptive prevalence remains low (32%), correct sexual and reproductive health (SRH) knowledge is only about 50%, and youth-friendly health services are scarce [6]. Consequently, roughly 25% of girls aged 15–19 become pregnant, and 97% of teen mothers drop out of school [7 8]. Qualitative research identified low contraceptive use as due to factors beyond knowledge and access, including concerns about misconceptions, fear of side effects and infertility, stigma, and embarrassment about accessing services that are not adolescent-friendly [9 10].

Since the 1994 International Conference on Population and Development, numerous SRH interventions have been implemented, mostly through schools, primary care facilities, and community stakeholders [11]. However, many have targeted married populations, reaching few unmarried adolescents, and have been limited in scale and reach. Consequently, many proved unsustainable and ineffective relative to investment [12–14]. Researchers have called for innovative interventions with robust designs—including implementation and cost-effectiveness analyses—to inform decisions on sustainability and scalability [11 15 16]. In Uganda, beyond abstinence-focused school education, services for sexually active adolescents have been delivered mainly through health facilities or community outreach [7 17]. However, access to these services remains limited because health facilities are unevenly available and outreach activities depend on donor funding [18]. Although multisectoral approaches have been promoted for two decades [19], an effective, sustainable, and nationwide model has yet to be established.

At the same time, mobile phone use in Uganda is widespread, with 63% of the population active users in early 2023 [20], and mobile money transaction volume grew 4.1-fold between 2015 and 2019 [4]. Our previous study found that most young people daily use mobile money vendors (MMVs) for money transfers, airtime purchases, and deposits [21], and that MMVs are ubiquitous nationwide.

Recognizing MMVs’ deep roots in the daily lives of young people, we developed an approach using these existing private, community-based channels as a new access point for contraceptive information and services—reaching underserved, unmarried adolescents whom conventional facility-based programs struggle to reach, while leveraging infrastructure that already operates nationwide with minimal additional investment to sustain. Having reported its effectiveness previously [21], this study assesses the feasibility of this MMV-based intervention using the Bowen feasibility framework.

## Methods

### Study design and setting

This feasibility study was embedded within a type II hybrid effectiveness-implementation study [22 23], with feasibility assessed using the Bowen feasibility framework [24]. The effectiveness component has been reported separately [21]; therefore, the present study focuses primarily on the feasibility and implementation of the MMV-based intervention, with feasibility assessed using the Bowen feasibility framework [24]. The intervention was conducted over four months from May 5, 2024 in the Busoga region. We reported the overall study design and effectiveness results in our publications [21 25]. The unit of intervention was MMV shops; the unit of analysis included MMVs, adolescents, and other stakeholders. Effectiveness was evaluated through a quantitative study of adolescent customers, while feasibility was assessed through MMVs’ focus group discussions (FGDs), key informant interviews (KIIs), a post-intervention MMV survey, MMVs’ service logs, and research team reports (Table 1).

**Table 1.** The Bowen feasibility framework and sources of verification.

| Area of focus | Definition | Source of verification |
|---|---|---|
| **Acceptability** | To what extent is a new program judged as suitable, satisfying, or attractive to program deliverers? To program recipients? | MMVs' log data<br>FGD with MMVs |
| **Demand** | To what extent is the use of the selected intervention in a defined intervention population (i.e., how much demand is likely to exist?) | Quantitative survey with adolescents<br>FGD with MMVs<br>Post-intervention MMV survey |
| **Implementation** | To what extent can the intervention be successfully delivered to intended participants? | FGD with MMVs<br>Reports of the weekly supervision and its observation |
| **Practicality** | To what extent can the intervention be delivered when resources, time, commitment, or some combination thereof are constrained in some way? | FGD with MMVs<br>Reports of the weekly supervision and its observation<br>Documents of the research team |
| **Adaptation** | Needs to modify the intervention contents or procedures to be appropriate in a new situation or with a different population. | FGD with MMVs<br>Reports of the weekly supervision and its observation |
| **Integration** | Needs for system changes to integrate the intervention into an existing infrastructure or program. | FGD with MMVs<br>Reports of the weekly supervision and its observation<br>KII with stakeholders |
| **Expansion** | The potential success of an already-successful intervention with a different population or in a different setting. | FGD with MMVs<br>Reports of the weekly supervision and its observation<br>KII with stakeholders |
| **Limited efficacy<sup>a</sup>** | Limitations of methods to assess the intervention effectiveness. | Quantitative survey with adolescents |

FGD, focus group discussion; KII, key informant interview; MMV, mobile money vendor.
<sup>a</sup> Limited efficacy is shown to present the complete framework; its findings are reported in the Limitations section rather than in the Results.

The Busoga region has one of the highest prevalence rates of adolescent pregnancy in Uganda [5]. Of the region’s four municipalities, we selected Iganga and Bugiri, which are located approximately 35 km from each other along the national highway connecting Kenya and Rwanda. Iganga and Bugiri municipalities have populations of 55,263 and 28,747, with 20.9% and 19.3% of those aged 10–17 years, respectively [4].

### Sampling

MMVs were purposively selected based on geographic distribution, ward population and characteristics, and operator profile (age, education, shop type and ownership, years in business, customer volume), since customers use multiple shops rather than being tied to one. Unmarried adolescents aged 15–19, resident in the target municipalities, with accessible phone numbers and no relocation plans within six months, were systematically recruited (every 35th customer) at the MMV sites at baseline and followed up at endline. The minimum sample size of 600 adolescents was calculated as described in the protocol [25].

### Intervention

The participating MMVs provided four services: oral information on SRH; information, education, and communication (IEC) materials on pregnancy, contraception, condom use, and sexually transmitted infections; free condoms (four per visit, wrapped in an envelope for discretion); and referral to health facilities. MMVs completed a two-day training before the intervention, and a mid-term workshop to reinforce SRH content and discuss challenges. MMVs kept a log on paper sheets; the research team conducted weekly supervision to monitor activities and supply condoms. The MMV shop was the unit of intervention; no shops were replaced or excluded during the intervention, and all 30 were retained in the analysis.

### Data collection

For the quantitative component, baseline (February–March 2024) and endline (September– November 2024) questionnaire surveys via data collectors were conducted with adolescents. The same adolescents were followed from baseline to endline. Respondents who were lost to follow-up or had missing responses to key variables at either round were excluded, and the analysis included only those with complete data at both baseline and endline. The questions covered sociodemographics, MMV usage, sexual activity, contraception, and SRH knowledge, capability, and motivation. The 30 participating MMVs kept daily logs of adolescent customers (aged 15– 19 years, verified through vendor knowledge and visual assessment) and of the services provided. At endline, we also conducted three FGDs with participating MMVs and eight KIIs with parents, teachers, and local health authorities. These explored perceptions, effectiveness, and scale-up potential; each session lasted 60–90 minutes and was audio-recorded. In February 2025, a post-intervention MMV survey, using a structured questionnaire, assessed business performance and mobile-company support. Weekly supervision reports were also used for analysis. The participant flow diagram is available elsewhere [21].

### Analysis

Effectiveness was evaluated using a quasi-experimental design, with condom use as the primary outcome and place of obtaining condoms as a secondary outcome. Analyses were conducted in Stata 18, with statistical significance set at p<0.05. Feasibility was assessed using Bowen’s feasibility framework, following the approach applied in previous evaluations of public health interventions [26]. Table 1 presents the Bowen framework with eight areas of focus, their definitions, and the sources of verification used in this study. The post-intervention MMV survey was analyzed descriptively. All FGDs and KIIs were audio-recorded in the local language, transcribed verbatim, translated into English, and analyzed manually using a deductive approach, with Bowen’s seven areas of focus serving as the coding framework. Combined findings for each area are reported in the Results section; the eighth area, limited efficacy, concerns how intervention effectiveness is assessed and is therefore addressed in the Limitations section. This manuscript was prepared with reference to the Transparent Reporting of Evaluations with Nonrandomized Designs (TREND) statement for the quantitative component and the Consolidated Criteria for Reporting Qualitative Research (COREQ) for the qualitative component.

### Ethics

This study was approved by the Uganda National Council for Science and Technology (SS2284ES), Uganda Christian University (UCUREC2023-710), and the Ogata Sadako Research Institute for Peace and Development, Japan International Cooperation Agency (JICA) (DI 202312220002), and registered with University Hospital Medical Information Network (UMIN000053332) on January 12, 2024. Written informed consent was obtained from all participants and, for those under 18, their parents or guardians.

## Results

The adolescent survey included 585 respondents in the intervention arm and 619 in the control arm at both baseline and endline (77 dropouts, 95.0% retention). Of these, 52.7% were male, the mean age was 17.8 (Standard deviation [SD]: ±1.2) years, 65.4% lived with parents, 58.5% visited MMVs daily. Detailed data are available in our previous paper [21]. As shown in Table 2, the post-intervention MMV survey had 30 participants. Of the 30, 73.3% of MMVs were female; the mean age was 26.4 (SD: ±3.7); 53.3% completed O-level (S4) and 26.7% had education above O-level (S5 or higher); 66.7% were single-owner, while the others were dual owners; Box-type (kiosk type) accounted for 73.3%. 90% of MMVs contracted with either one or two mobile carrier companies (Airtel, MTN), and 87% of them received daily supervision from company supervisors. For the FGDs with MMVs after the intervention, 26 MMVs participated, of whom 20 were female (Table 2). KII participants are summarized in Table 2. As most participants were not aware of the intervention, their comments reflected hypothetical views rather than direct observations. The qualitative analysis, therefore, relied primarily on the MMV FGDs. The seven areas of focus assessed are described below; notable quotes are numbered for reference and listed in Table 3.

**Table 2.** Characteristics of study participants.

| **Characteristic** | **Post-intervention MMV survey (n = 30)** | **FGD with MMVs (n = 26)** | **KII (n = 8)** |
|---|---|---|---|
|  | **n (%)** | **n (%)** | **n (%)** |
| **Sex** |  |  |  |
| Female | 22 (73.3) | 20 (76.9) | 3 (37.5) |
| Male | 8 (26.7) | 6 (23.1) | 5 (62.5) |
| **Age (years)<sup>a</sup>** |  |  |  |
| Mean (SD) | 26.4 (3.7) | 26.4 (3.9) | 42.1 (8.7) |
| **Age group** |  |  |  |
| ≤24 | 10 (33.3) | 9 (34.6) | 0 (0.0) |
| 25–29 | 10 (33.3) | 7 (26.9) | 0 (0.0) |
| 30-39 | 10 (33.3) | 10 (38.5) | 3 (37.5) |
| 40–49 | 0 (0.0) | 0 (0.0) | 4 (50.0) |
| ≥50 | 0 (0.0) | 0 (0.0) | 1 (12.5) |
| **Education completed** |  |  |  |
| Below O-level (P7–S3)<sup>b</sup> | 6 (20.0) | 6 (23.1) | 2 (25.0) |
| O-level (S4) | 16 (53.3) | 16 (61.5) | 0 (0.0) |
| Above O-level (S5 or higher) | 8 (26.7) | 4 (15.4) | 6 (75.0) |
| **Type of MMV** |  |  |  |
| Box (Kiosk) | 22 (73.3) | — | — |
| Mix shop<sup>c</sup> | 5 (16.7) | — | — |
| Veranda | 2 (6.7) | — | — |
| Umbrella | 1 (3.3) | — | — |
| **MMV ownership** |  |  |  |
| Single | 20 (66.7) | — | — |
| Dual | 10 (33.3) | — | — |
| **Type of key informant** |  |  |  |
| Health authority | — | — | 2 (25.0) |
| School teacher | — | — | 2 (25.0) |
| Parent | — | — | 4 (50.0) |

MMV, mobile money vendor; FGD, focus group discussion; KII, key informant interview; SD, standard deviation.
<sup>a</sup> Survey ages are from the vendor list as of the post-intervention survey in February 2025.
<sup>b</sup> For education, one KII parent completed P6.
<sup>c</sup> “Mix Shop” indicates MMVs who were running small commodity shops and selling daily commodities or stationery.
— = not applicable or not collected. Percentages may not sum to 100 due to rounding.

**Table 3.** Notable quotes by identified sub-theme.

| Area of focus | Sub-theme | Narrative |
|---|---|---|
| **Acceptability** | Satisfaction with adolescents | “Most of the teenagers were very happy and they are very thankful because initially they were buying those things from clinics, but now, they could get them for free with no transport costs since services were extended nearer to them.” (F, 25–29) #1<br>“At first the teenagers were scared thinking that I might share what they told me with their parents, because they could see me talking with their mothers. But it wasn't the case, so they are very grateful for this service.” (F, 20–24) #2 |
|  | Adults’ reaction | “Even some parents used to request some condoms to take for their kids and IEC materials to take to their girls to read through.” (F, 25–29) #3<br>“Sometimes a parent can come with her child and tells her child to sit there then the parent makes a sign for me to talk to her child. Then another parent came with her child who has just aborted so… I also explained to her the dangers of aborting.” (F, 20–24) #4<br>“Teachers did not see any importance of the research at first, but when we gave them the brochures [IEC materials], they read through and afterwards many of them took their fellow teachers and students.” (F, 25–29) #5 |
|  | Conflicts | “I have a friend of a pastor who came to pray for my business, and he saw those IEC materials and asked me what they were. He told me that family planning is very bad according to Christians because you are trying to kill the babies inside the stomach; therefore, our friendship reduced.” (F, 30–34) #6 |
|  | Low referral | “When you send her for referral, she asks you to escort her, so what I do is: I get the referral and give it to the doctor myself.” (F, 25–29) #7<br>“I referred a girl to Islamic but the girl came back and told me that she is tired of going there because every when she goes there, they tell her to first to wait. So my request is that… when we send their referrals, they go straight to that health workers and [get services].” (F, 30–34) #8 |
| **Demand** | High adolescents’ demands | “After teaching these people how to use these family planning methods and since they used to fear to face the health workers, and they were free with us, the mobile money attendants, they got more information on how to use condoms and all those myths about condom, like the condom may remain in you after use, are false. Therefore, people keep on coming asking for condoms.” (M, 30–34) #9<br>“Because most of these girls in the community used to get pregnant, but in this period of four months, I am not seeing any teenage girls pregnant. Even the health workers are surprised that the rate of abortion has reduced.” (F, 30–34) #10 |
|  | Vendors’ benefits | “Most of these teenagers became my friends because when I teach them about family planning methods, they go back and told their sisters and brothers about our services. So, they come for a deposit like one thousand shillings or water for five hundred shillings, then ask for condoms.” (F, 20–24) #11<br>“It has helped me because my customers increased in number because we teenagers that stay in **** [outside of Iganga] but they move from those ends and come here.” (F, 20–24) #12<br>“My neighbor [MMV] was not around the first time they [research team] registered us. So… she asked where I got them… I told her that if I get another chance, I will tell them to give you. But they all welcomed the service.” (F, 25–29) #13 |
| **Implementation** | Smooth implementation | “Work has been moving smoothly in these four months we have taken, while working for [our business].” (F, 25–29) #14 |
|  | Discreet packaging | “These teenagers were very happy when you gave us this wrapping because no one can realize what he/ she is taking in that envelope.”(F, 30–34) #15 |
|  | Negative reactions | “They [adults] used to ask, ‘Why are you interviewing children of fifteen years, they know nothing?’ I replied to them by saying that those children you see know everything and have a lot of experience and they could understand whatever we were talking to them.” (F, 25–29) #16<br>“The chairman reacted so badly because he even called [name of the research team] he wanted them to first go to his office for approval… Maybe he wanted some money from them.” (F, 30–34) #17 |
|  | Promoting factors | “Girls ask me if those family planning methods don't spoil their uterus. At that time, I also didn't have enough knowledge about that. But after I got a second training, I got more knowledge and I went back and taught them.” (M, 25–29) #18 |
| **Practicality** | No significant obstacles to business | “It took me like fifteen minutes. Although some could ask just for the condom and the brochures to go and read by themselves, some wanted me to first explain them.” (F, 25–29) #19<br>“It depended on [teens], some could take a long time, especially for IEC, for which someone needed clarification, so it could take me a longer time to explain.” (F, 20–24) #20 |
| **Adaptation** | Compatibility with the mobile money business | “My humble request is that for easy identification we should at least be given T-shirts and identification cards so that we work with confidence.” (F, 30–34) #21<br>“They commonly complained that the condoms from government of Uganda so they wanted to change them.” (M, 20–24) #22<br>“I think to expand our services, they should add more items like sanitary pads, not only condoms because right now… boys that have a big turn up.” (F, 25–29) #23<br>“I just want to thank you very much for my salary.” (F, 20–24) #24 |
| **Integration** | Integration with public systems | “…we can look at integrating it into the system… by involving other stakeholders, the policy makers so that they come to know… the minister of health that is at national level then at the district level this is the district leadership including the councilors then from the district we can escalate it down to the sub counties… These vendors or the mobile money vendors can be brought on board because the government is trying to roll out community extension health workers… we can consider them as part of our system or VHT system…” (KII, health authority) #25<br>“…maybe you could work with these companies and some message comes on these phones… so that somebody has got that kind of permanent information.” (KII, teacher, M, 40s) #26<br>“I think the saloon people….can give you time as he is shaving your hair and he explains to you…other places of social gatherings like cinema halls and even boreholes and wells, could even save more lives.” (KII, teacher, M, 40s) #27 |
| **Expansion** | A widely spread business network | “…these people are widespread now… you can go to any area, even in rural… these mobile money people are there… it is a good business and it reaches people directly…” (KII, teacher, M, 40s) #28 |
|  | High demand in rural areas | “Even in villages children are far from the hospitals and those children cannot go to the hospitals to get the condoms and they are even shy. If that program is extended to the villages it can work.” (M, 25–29) #29<br>“We can approach nearby areas so that they can get services like this, so that we can teach our untrained colleagues these services by us, the trained people. I think it should be like that…” (F, 30–34) #30 |

F, female; M, male;  IEC, information, education and communication; KII, key informant interview; VHT, village health team.
For MMVs, age is shown as a 5-year age group; for key informants, as a 10-year age group.
The numbers preceded by the # symbol indicate the references in the text. Words in square brackets were added by the authors for clarity; \*\*\*\* indicates a name withheld.

### 1) Acceptability

Overall, the intervention was well accepted by both adolescents and MMVs, with high satisfaction. Over the four-month intervention, participating MMVs recorded 104,849 adolescent customer visits; of that oral SRH information was provided at 54,365, IEC materials at 39,141, and referrals at 210 cases. Condoms were provided at 21,056 visits, 83% to males. Customer numbers and condom provision remained stable across the intervention, with peaks during school holidays in late May and late August (Figure 1).

**Figure 1.**
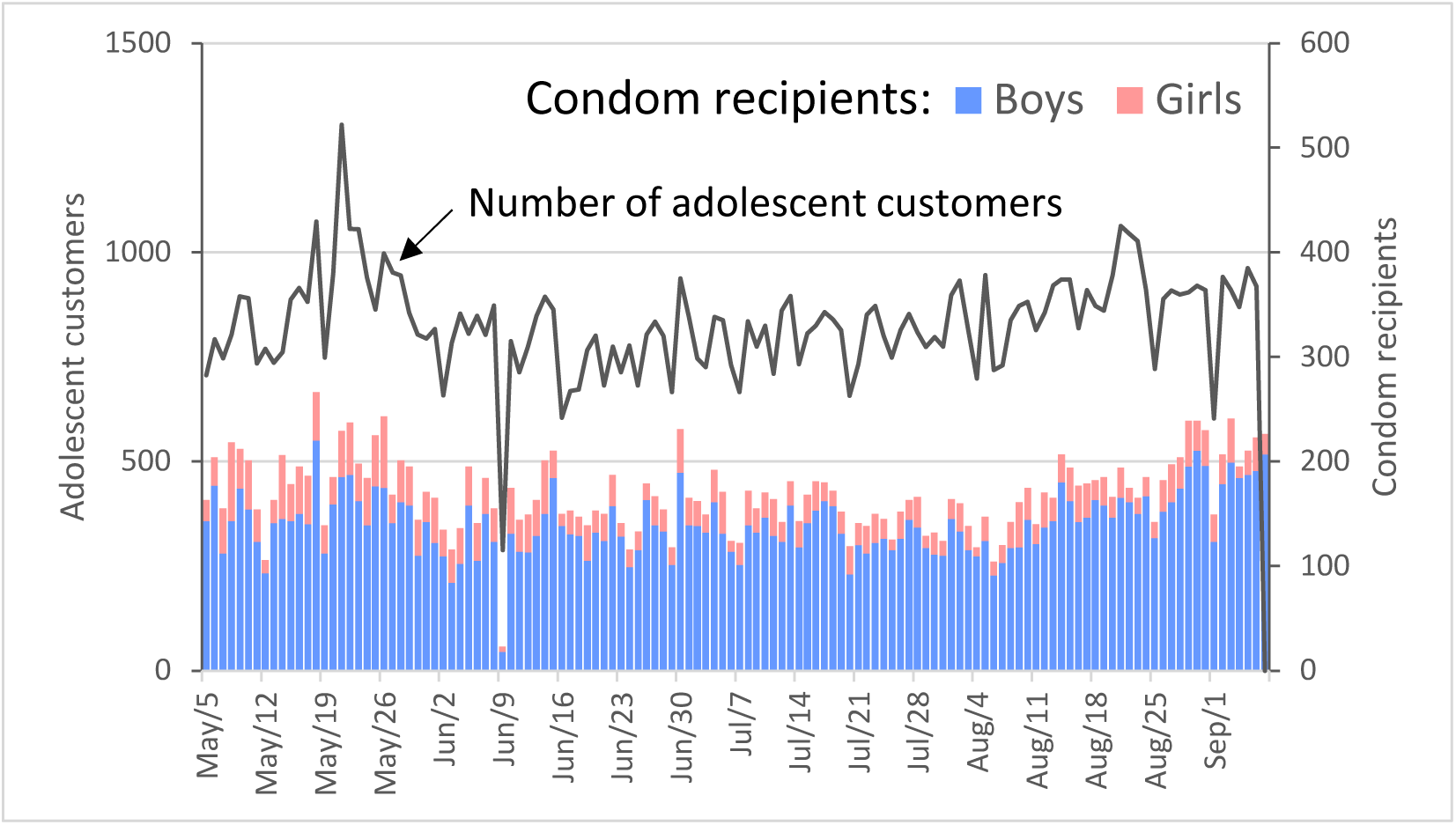
Daily numbers of adolescent customers and condom recipients at the 30 participating MMVs Data are from MMV service logs. The line graph shows the daily number of adolescent customers, while the bar graph shows the number of adolescents who received condoms (blue represents males, and pink represents females). The significant drop on June 9 was because the research team convened the MMVs for a training session.

Most adolescents appreciated obtaining these services free of charge at community-based MMVs, which were familiar neighborhood fixtures. Some MMVs initially found it difficult to discuss SRH with adolescent girls, who were shy and feared disclosure to parents, but MMVs developed skills to deliver such sensitive issues and assured confidentiality. MMVs attributed the high acceptance to their closeness in age to adolescents, which fostered familiarity and trust, and to their convenience (#1-2).

Some parents, teachers, and religious leaders showed initial resistance, believing the intervention would encourage sexual activity. This resistance largely dissipated over time—parents whose children faced sexual risk became active supporters, and skeptical teachers changed their views after seeing the IEC materials (#3-5). However, a few religious leaders continued to oppose condom provision (#6).

The low referral rate (210 cases) reflected demand- and supply-side barriers: adolescent girls were often too shy to attend facilities alone (#7) or feared confidentiality breaches, while on the supply side, the referral system was not adequately prepared—some health care workers (HCWs) were unaware of the intervention or declined the added task without incentive, and formal integration into the official referral system had not been arranged. Some HCWs also failed to direct adolescents appropriately or questioned their need for contraceptives (#7-8).

### 2) Demand

Demand was reflected in the substantial uptake of services described above. Consistent with this observed demand, the previously reported effectiveness analysis showed a substantial shift in the source of condoms toward MMVs: the proportion of participants obtaining condoms from MMVs increased from 0% at baseline to 64.6% at endline in the intervention arm. Condom use increased by 10.4 percentage points among the intervention arm and other effectiveness outcomes have been reported in detail elsewhere [21]. MMVs also described how providing information and addressing misconceptions about condoms, along with adolescents’ comfort in interacting with MMVs, appeared to encourage adolescents to return to request condoms (#9). Several MMVs also shared a perception, based on accounts from other MMVs and HCWs, that unintended pregnancies and abortions among adolescent girls may have declined (#10), although this study did not measure these outcomes.

According to the post-intervention MMV survey (n = 30), average daily numbers of adolescent customers rose to 1.64-fold the pre-intervention level during the intervention, and average daily sales rose 1.50-fold, remaining 1.21-fold higher after the intervention ended (Figure 2). Many reported that once adolescents got services at the MMVs, they brought back their brothers/sisters or friends (#11). Even young people outside the intervention areas came to seek the services (#12). As the intervention progressed, relationships with community members improved, the community showed appreciation for the intervention, and the majority of MMVs were highly motivated to continue it. In addition, other MMVs surrounding the participating ones were also eager to join (#13).

**Figure 2.**
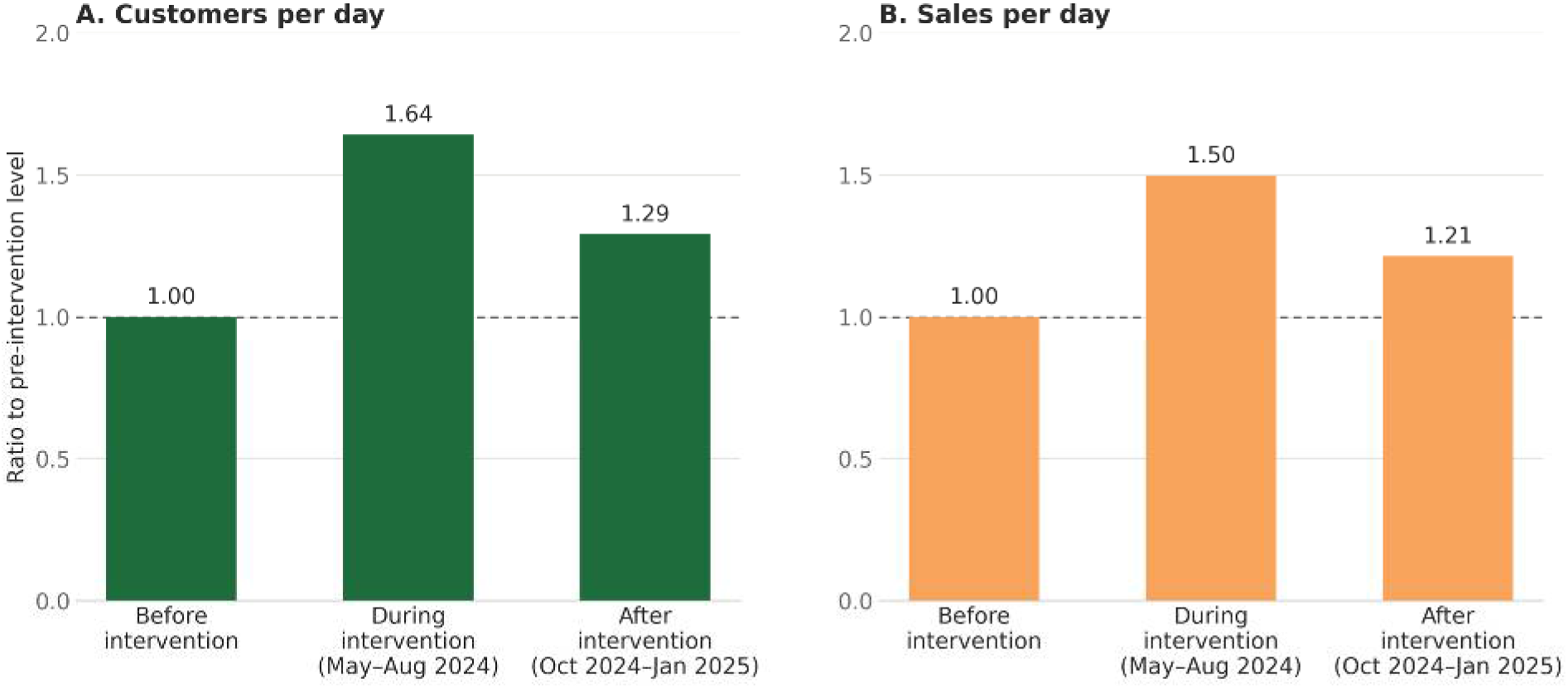
Changes in (A) customers per day and (B) sales per day of participating MMVs before, during, and after the intervention (n = 30) Data are from the post-intervention MMV survey (February 2025). Values are ratios of group means to the pre-intervention mean. The dashed line shows the pre-intervention level.

### 3) Implementation

All 30 MMV shops completed the four-month intervention. In two shops, the original attendant was replaced by another person at the same location within the first 10 days and stayed until the end, owing to illness or a misunderstanding of the role; the new attendants were trained on site by the research team. Most reported no significant implementation challenges or conflicts with their business (#14). Wrapping the condoms in an envelope was highly appreciated by recipients (#15). Although some faced initial difficulties, these diminished over time. The most significant challenge, as described earlier, came from local adults, including parents, school teachers, and religious leaders, some of whom initially opposed the provision of SRH information and condoms (#16), believing this would encourage adolescent sexual activity. A small number of local politicians expressed dissatisfaction that they had not been briefed before the intervention commenced (#17). These were addressed through MMVs’ continuous communication using posters and IEC materials, along with support from the research team, to foster understanding of the study’s purpose and the importance of educating adolescents on safe sex and providing condoms to prevent unintended pregnancy and sexually transmitted infections.

Many MMVs stated that the research team’s on-site supervision was effective in implementation, which included clarifying points of confusion, helping resolve conflicts with influential local people, and providing supplies on time. In addition, timely training at the beginning and mid-term was also effective in providing necessary SRH information and sharing experiences among peer MMVs (#18; weekly supervision reports).

### 4) Practicality

Time required for implementing the intervention was not a major constraint for most MMVs. The first contact typically took about 15 minutes—explaining the intervention’s purpose, obtaining acceptance, providing SRH information and distributing condoms (#19). Some adolescents requested deeper explanations of IEC materials, extending this time (#20). However, after the second or third visit, interactions were brief, as adolescents simply picked up condoms and left. The research team noted that a few MMVs initially struggled to maintain daily logs. Continuous on-site supervision resolved this within a few weeks (weekly supervision reports). The estimated implementation cost over four months—covering training, supervision, IEC materials, vendor compensation, and administration, but excluding condoms—was approximately 14,650 United States dollar [USD], or 0.70 USD per condom recipient (including SRH information) (research team documents). If the intervention were run by a public agency, with administration absorbed into routine operations, IEC materials supplied through existing government stock, and no vendor compensation, only training and supervision costs would remain: approximately 4,110 USD, or 0.20 USD per condom recipient.

### 5) Adaptation

MMVs identified several areas for further adaptation. Many requested identification, such as ID cards or T-shirts, to signal their official participation (#21). Because MMV selection criteria were set narrowly for research purposes, wider scale-up would require broader and more transparent criteria, potentially open to anyone with basic capability and motivation to serve the community. According to MMVs, adolescents’ main complaints about the condoms supplied by the Ministry of Health concerned their odor and size, and they requested better-quality alternatives (#22). MMVs also noted that girls often perceived the intervention as male-oriented and wanted a wider range of services, including other contraceptive methods, sanitary pads, and diapers (#23). Some MMVs regarded the monthly compensation as a salary, which may have shaped their reasons for participating in ways the intervention did not intend (#24).

### 6) Integration

A local health authority suggested integrating the approach with the Ministry of Health system, including a shared health information system to capture service provision and commodity supply, potentially using a community volunteer mechanism (#25). One teacher proposed mobilizing additional community channels, such as schools, cinema halls, wells, and hair salons, and partnering with mobile carrier companies to deliver SRH messages alongside condom transactions (#26-27).

### 7) Expansion

The majority of MMVs and stakeholders indicated that this approach could be applied to other areas. Some emphasized it should be applied to rural villages where there is a greater lack of SRH information and services for unmarried adolescents (#28-29). Many of the MMVs also showed eagerness to be involved in such expansion since they became confident in contributing to social services (#30). One school teacher framed the underlying rationale for scaling: MMVs are geographically embedded and trusted, which makes the approach portable (#28). A health authority emphasized the potential for a system-level scale-up, with the public health structure, for nationwide expansion (#25).

### 8) Limited efficacy

Statistical power was lower than expected because the assumed contraceptive use rate used to calculate the sample size was lower than the expectation in the study area. Additionally, other programs providing condoms might be implemented in the control area.

## Discussion

This study makes three contributions: we developed an innovative approach in Uganda that uses an existing private-sector channel; the approach reached previously hard-to-reach unmarried adolescents and was accompanied by increased condom use; and we evaluated this social implementation experiment scientifically, yielding practical recommendations for scale-up.

The MMV-based approach recorded over 100,000 adolescent customer visits in four months, with SRH information provided at more than half of these visits, while condom use in the intervention arm rose by approximately 10 percentage points—over 60% of condom users now obtaining them from MMVs rather than public institutions. MMVs also benefited from increased customers and sales and reported strong community trust and intent to continue.

An important finding was the marked gender difference in service uptake: 83% of adolescents who obtained condoms from MMVs were male. This is consistent with the previously reported effectiveness analysis, in which improvements in condom use and some measures of contraceptive motivation were more pronounced among boys [21]. Qualitative findings provided some context for this gender difference: MMVs reported higher service uptake among boys and suggested broadening the services beyond condoms, for example, by including other contraceptive methods and sanitary pads, to better respond to girls’ needs. These findings suggest the need to consider how the intervention could be adapted to better engage adolescent girls.

The Bowen framework identified several practical recommendations for further implementation and potential scale-up: selecting MMVs who can reliably maintain confidentiality; advance notification to community leaders and parents; strengthening confidentiality and service diversification for girls as mentioned above; collaboration with other community-based outlets (e.g., hair salons, wells); improving adolescent-friendly referral services; and integrating with existing public health mechanisms and schools. MMVs reported that tailored training and close supportive supervision facilitated smooth implementation, and that printed materials supported acceptance among adolescents and community stakeholders.

This approach may also address factors contributing to unintended pregnancies—such as a lack of knowledge about contraception, poor access to contraceptives, a lack of parental involvement, and the absence of accessible channels within the community, as the previous study identified [3]. It also encountered some initial opposition to providing family planning information and services to adolescents, consistent with previously reported sociocultural barriers to adolescent contraceptive use and access to sexual and reproductive health information and services in Uganda [10 27 28]. At the same time, some community members became more supportive after receiving information about the intervention. These findings suggest that by engaging community members and increasing awareness of adolescent SRH needs, the approach may contribute to reducing stigma surrounding contraceptive use among girls in the community.

Conventional adolescent SRH interventions have relied largely on schools, health facilities, and outreach, often collapsing once donor funding ends [7 17]. Despite repeated calls for innovative solutions, few have addressed this sustainability gap [11]. Our approach is novel in leveraging existing private-sector infrastructure to prioritize sustainability—offering a concrete model where public–private partnership has long been cited as important to Uganda’s SRH policy but never operationalized [29]. As infrastructure operating under a mobile network’s oversight, MMVs already benefit from guaranteed accountability and stability (e.g., Airtel Uganda’s daily franchise supervision), reinforcing their suitability as complementary public-service channels.

This study had several limitations. First, the assumed condom use rate used for sample-size calculation was lower than the actual rate observed (30% assumed vs. 67% actual), leaving the study underpowered; future studies in Uganda should allow a wider margin when local statistics are imprecise. Second, despite selecting control areas without other SRH programs, we later found an unrelated donor-funded family planning program operating there, reflecting the difficulty of finding a truly untouched control area amid sporadic regional health programming. Third, budget constraints limited the intervention to four months; assessing sustainability may require follow-up around one year while MMVs continue voluntarily. Fourth, MMVs’ compensation may have partly sustained motivation; future studies should test whether business benefits and civic motivation alone can sustain participation. Fifth, much of the qualitative research was based on MMVs’ FGDs. This was because many of the KII participants had no direct experience with the intervention. Consequently, a limitation of this study is that the perspectives of adolescents, the intended primary beneficiaries, were not captured directly through qualitative methods.

## Conclusion

Despite these constraints, this study suggests that existing MMVs can serve as a private-sector channel for providing condoms and SRH information to hard-to-access unmarried adolescents in resource-limited settings. The approach was feasible during the four-month supported implementation period and was accompanied by increased condom use among adolescents in the study area. However, weak referral linkages and lower service uptake among girls should be addressed before larger-scale implementation in Uganda, which also requires further evaluation of sustainability and costs at scale. Similar models could be piloted in neighboring Sub-Saharan African countries facing comparable demographic and social challenges.

## Data Availability

All data produced in the present study are available upon reasonable request to the authors.

De-identified data supporting the findings of this study will be shared through the University Hospital Medical Information Network (UMIN; registration number UMIN000053332). In addition, de-identified data are available from the corresponding author upon reasonable request.

https://www.umin.ac.jp/ctr/

## Sources of funding and acknowledgments of support and assistance

This work was supported by JSPS KAKENHI (grant number JP23K16346) from Grants-in-Aid for Scientific Research from the Ministry of Education, Culture, Sports, Science and Technology of Japan, which had no role in study design, analysis, or the decision to submit. We would like to express our gratitude to all study participants and the Ministry of Health for their invaluable support. We are also grateful to the JICA Ogata Sadako Research Institute for Peace and Development for providing research funding and for its support in implementing this study. We also extend our heartfelt thanks to all members of the research team of the Busoga Health Forum, whose efforts facilitated the implementation of this study, including the development of a network involving various stakeholders and healthcare facilities.

## Disclosure of potential conflicts

The authors have no conflicts of interest to declare.

## Clinical trials registry

This study was registered with the University Hospital Medical Information Network in Japan (UMIN000053332) on January 12, 2024.

## Authors’ contributions

MK, MS, and SH were involved in the conception and design of the study. MK and PK were responsible for field research implementation. SH and PW provided technical advice on the overall methodology, including sampling. MK drafted the manuscript. PW supervised the study and was involved in critical revision of the manuscript. MS, SH, and PW provided technical comments on the manuscript draft. MK was responsible for funding acquisition. All authors approved the final manuscript and the decision to submit it for publication.

## Reporting guidelines

This manuscript was prepared with reference to the TREND statement for the quantitative component and the COREQ checklist for the qualitative component.

## List of abbreviations

COREQ: Consolidated Criteria for Reporting Qualitative Research
FGD: Focus group discussion
HCW: Health care worker
IEC: Information, education, and communication
JICA: Japan International Cooperation Agency
KII: Key informant interview
MMV: Mobile money vendor
SD: Standard deviation
SRH: Sexual and reproductive health
UMIN: University Hospital Medical Information Network
TREND: Transparent Reporting of Evaluations with Nonrandomized Designs
USD: United States dollar

## Notes

### Competing Interest Statement

The authors have declared no competing interest.

### Clinical Trial

UMIN000053332

### Clinical Protocols

https://bmjopen.bmj.com/content/14/4/e084539

### Author Declarations

The ethics review committees of the Uganda National Council for Science and Technology (SS2284ES), the Uganda Christian University (UCUREC2023-710), and Ogata Sadako Research Institute for Peace and Development, Japan International Cooperation Agency (JICA (DI) 202312220002) approved the study.

